# Pilot randomized controlled trial of acetylsalicylic acid to reduce cerebral microembolism in Chagas heart failure

**DOI:** 10.1101/2024.10.09.24314743

**Authors:** Renan Carvalho Castello-Branco, Cárita Victoria Carvalho de Santana, Victor L. P. P. Botelho, Paulo R. S. P. deSousa, Maria C.P. Nunes, Karen L. Furie, Jamary Oliveira-Filho

**Author notes:** Corresponding author: Jamary Oliveira-Filho, Neurologist and Post-Graduate Program in Health Sciences (PPgCS), Hospital Universitario Professor Edgard Santos, Federal University of Bahia, Brazil.

## Abstract

**Background:** Chagas disease is an important cause of heart failure (HF) and stroke, affecting over six million people. High intensity transient signals (HITS) are detected on transcranial Doppler (TCD) in patients with Chagas disease, but the effect of antithrombotic treatment on HITS is unknown. The aim of this study was to evaluate whether acetylsalicylic acid (ASA) reduces the frequency and number of HITS in patients with chagasic HF.

**Methods:** Proof-of-principle pilot prospective, randomized, open and blinded endpoint clinical trial (PROBE), where patients with both Chagas and HITS were randomized 2:1 to ASA 300 mg for seven days and standard HF treatment or standard HF treatment alone (control group). The primary outcome was the proportion of HITS after one week, analyzed using chi-squared test.

**Results:** 373 patients with HF were evaluated, HITS occurred in 22/190 (12%) chagasic patients and in 16/183 (8%) non-chagasic patients (p=0.531). Twelve of the 22 (54%) chagasic patients were randomized to treatment with ASA (n = 8) or without ASA (n = 4). Two patients in the control group (50%) persisted with HITS after 7 days of treatment, compared to none in the ASA group, p=0.028. Median number of HITS decreased from 3.5 to 0 with ASA (p=0.012) and 4.0 to 0.5 in the control group (p=0.095), with no significant between-group difference (p=0.262). No adverse events were reported.

**Conclusion:** In this pilot clinical trial, ASA reduced the proportion of HITS in patients with Chagas disease HF.

## Introduction

Chagas disease affects over 6 million people worldwide, mainly in Latin American countries; however, migration patterns have increased infected individuals in Europe, Japan, Australia, Canada and the United States.^1^ Victims are often infected as children and up to a third eventually develop cardiomyopathy for the rest of their lives, increasing the risk of sudden death from arrhythmias and cerebral infarction.^2, 3^

The establishment of non-invasive methods to stratify the risk of stroke in patients with Chagas disease is a priority. Additionally, it is important to define alternative outcomes to test new treatments for stroke prevention in this population. Transcranial Doppler (TCD) is a non-invasive and safe method that allows the identification of microembolism in the cerebral circulation in real time. Inflammation and secondary activation of the hemostatic system is thought to increase the risk of thrombus formation and subsequent embolic stroke, especially in the presence of structural myocardial damage such as dilated cardiomyopathy.^4, 5^ The microembolic events are also called high intensity transient signals (HITS), visualized on TCD as high intensity signals (usually> 3dB above its basal value), unidirectional, transient and with a specific sound.^6^

In patients with heart failure (HF), HITS occur more frequently in Chagas when compared to non-Chagas patients^7^, but it is unknown whether this increased risk can be modified by anti-thrombotic medications. In this sense, the aim of the study was to test the effectiveness of acetylsalicylic acid (ASA) in reducing the rate of HITS in patients with Chagas-associated heart failure.

## Methods

This was a pilot proof-of-principle prospective, randomized, open-label, blinded clinical (PROBE) trial. Inclusion criteria were age above 18 years of age with HF defined according to Framingham clinical criteria formed by signs (progressive edema of the lower limbs and hepatomegaly not attributable to other diseases) and symptoms (dyspnea on exertion and paroxysmal nocturnal dyspnea) and classified according to the New York Heart Association functional class.^8, 9^ In addition, patients with Chagas disease had two positive serological tests (ELISA, immunofluorescence or hemagglutination assays). Patients with a history of stroke or atrial fibrillation related to cardiomyopathy were excluded.

Exclusion criteria were designed to avoid confounding variables for transcranial Doppler HITS detection, such as co-morbidities and use of oral anticoagulants. Patients with a history of untreated malignant neoplasm (except localized neoplasm of the skin), ischemic cerebrovascular disease (determined using the Questionnaire for Verifying Stroke-Free Status),^10^ chronic dialysis renal failure or end-stage liver failure were excluded. Patients with HITS detected in the TCD were screened for exclusion criteria for using ASA.

Patients screened following the inclusion / exclusion criteria underwent TCD monitoring with a helmet. A single investigator performed all tests, blinded to all clinical data. The middle cerebral artery was monitored continuously unilaterally for one hour looking for the occurrence of HITS. Chagasic patients with HITS detected during monitoring were randomly allocated 2:1 to treatment with ASA 300mg for 7 days and their usual medications for HF treatment; or their medications for HF treatment alone. These patients performed a second TCD after seven days verify the effectiveness of ASA in reducing HITS with an investigator blinded to treatment allocation.

### Statistical analysis

The clinical data collected in Brazil was entered into a Research Electronic Data Capture (REDCap) database and analyzed using STATA version 14.1. Demographic and clinical characteristics were described as absolute and relative frequencies for categorical variables and mean/standard deviation or median/interquartile range for continuous variables, depending on its distribution. Normality of continuous variables were defined based on Kolmogorov-Smirnov test. Based on our preliminary data on the frequency of HITS in cases and controls (14% and 2%, respectively), we expected to recruit 37 chagasic patients with HITS. Considering a 15% refusal rate, we expected to randomize 30 patients for ASA or control treatment. With this sample size, we expected to have 80% power to detect a minimum reduction of 58% in the proportion of HITS in the ASA group compared to 10% in the control group. Unfortunately, the study was interrupted prematurely due to lack of funding and higher than expected refusal rates.

To compare the proportion of HITS between groups, we used chi-squared test with Yates correction. As a secondary analysis, we compared the median number of emboli using Mann Whitney U test. To compare the proportion of HITS between Chagas and non-Chagas groups we used logistic regression analysis to control for confounding factors.

The data that support the findings of this study are available from the corresponding author upon reasonable request. The study was approved on February 13, 2014 by the ethics committee of Professor Edgard Santos University Hospital - Federal University of Bahia, Brazil and all patients signed informed consent. This clinical trial and its analysis plan were registered at www.clinicaltrials.gov under number NCT01650792.

## Results

Of the patients included in the study, 373 underwent TCD, 190 (51%) with HF Chagas disease, median age 59 (interquartile range 35-65), 313 (83.9%) afrodescendants, 123 (65%) females. Most frequent cerebrovascular risk factor was hypertension, present in 116 (62%) patients. Median left ventricular ejection fraction was 42% (interquartile range 28-62). Chagas patients less frequently had comorbidities associated with other HF etiologies such as coronary artery disease, hypertension and diabetes. Table 1 shows the baseline characteristics of both populations.

**Table 1.**
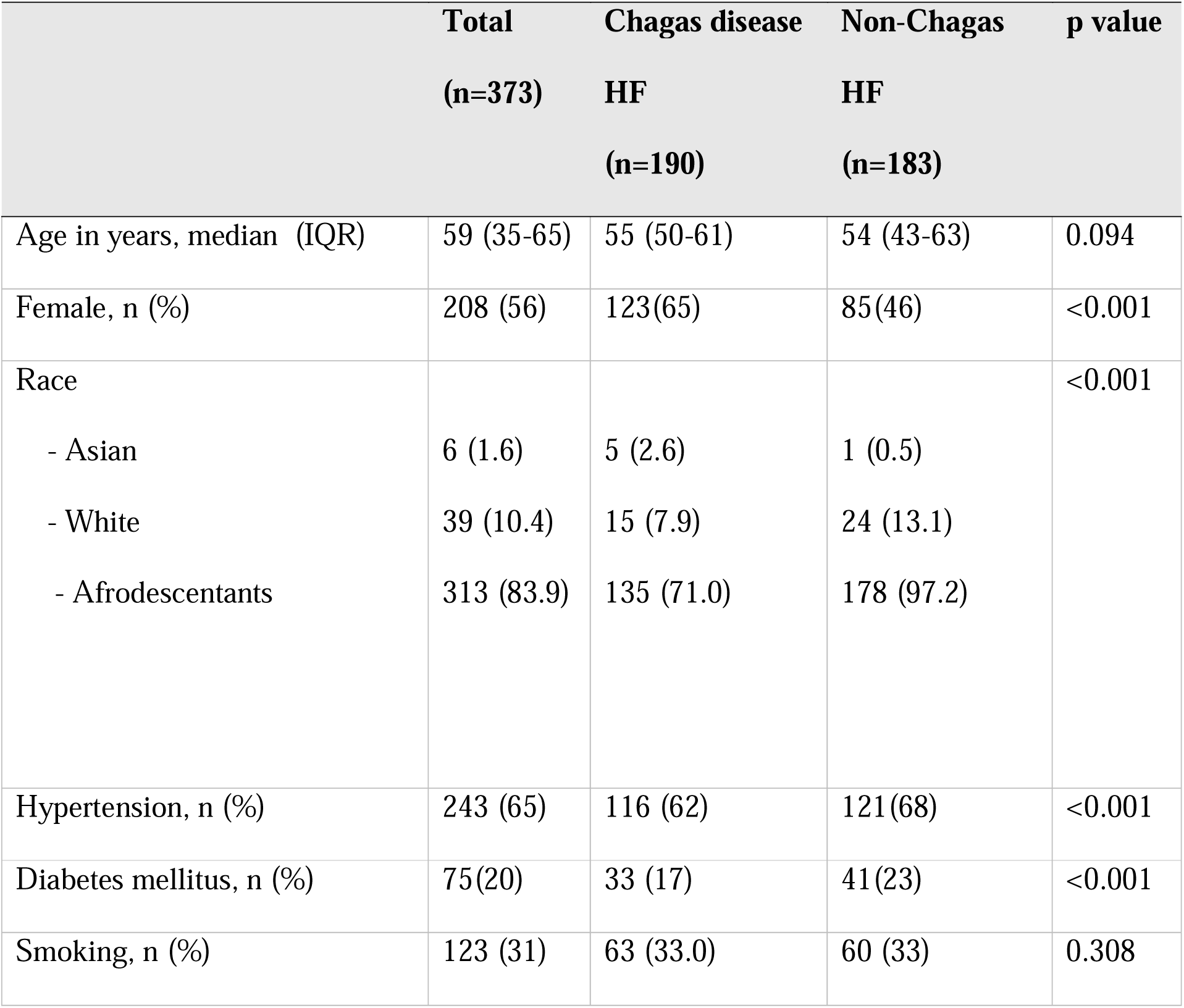
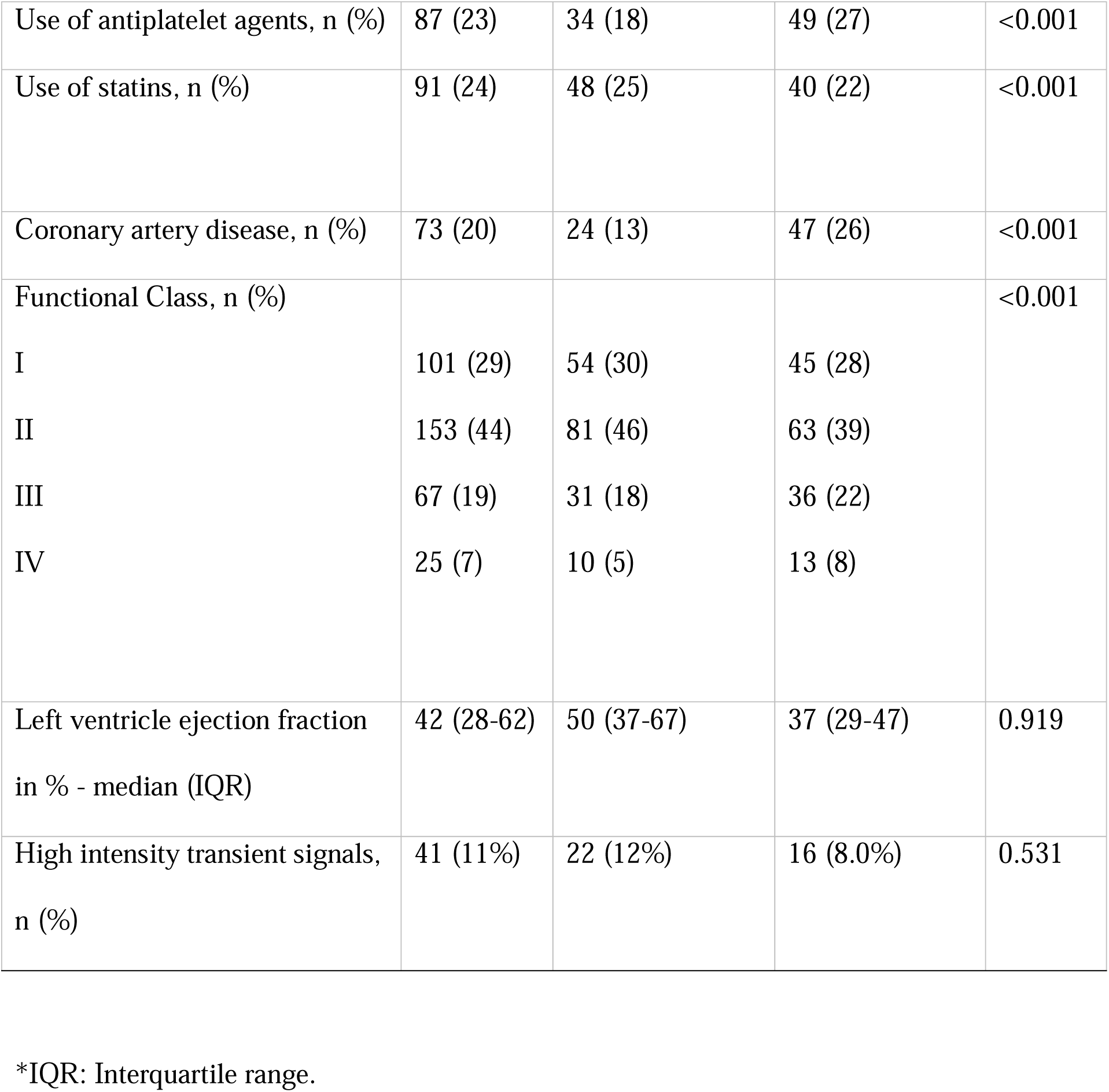
Baseline characteristics of patients with heart failure (HF) of Chagasic vs Non-Chagasic etiology.

Of the patients who underwent TCD, 41 (11%) patients presented with HITS, observed in 22/190 (12%) of chagasic patients and in 16/183 (8.0%) of non-chagasic patients (p=0.531). In the multivariable analysis adjusted for age, sex and left ventricle ejection fraction, Chagas disease was not associated with HITS (odds ratio = 1.33; 95% confidence interval = 0.60 – 2.95).

Twelve of the 22 patients (54.5%) with Chagas disease were randomized to treatment with ASA (n=8) or a control group (n=4). Characteristics of both groups were similar (Table 2). The remaining patients refused consent mostly due to logistical issues (e.g., lived in a city far from the stroke center). After seven days, two patients in the control group (50%) persisted with HITS, while none in the ASA group persisted with HITS, p=0.028.

**Table 2.**
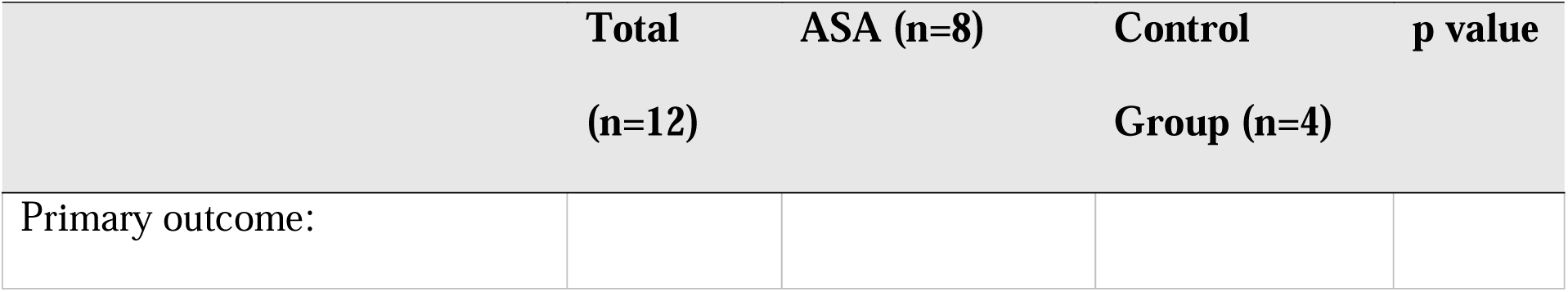
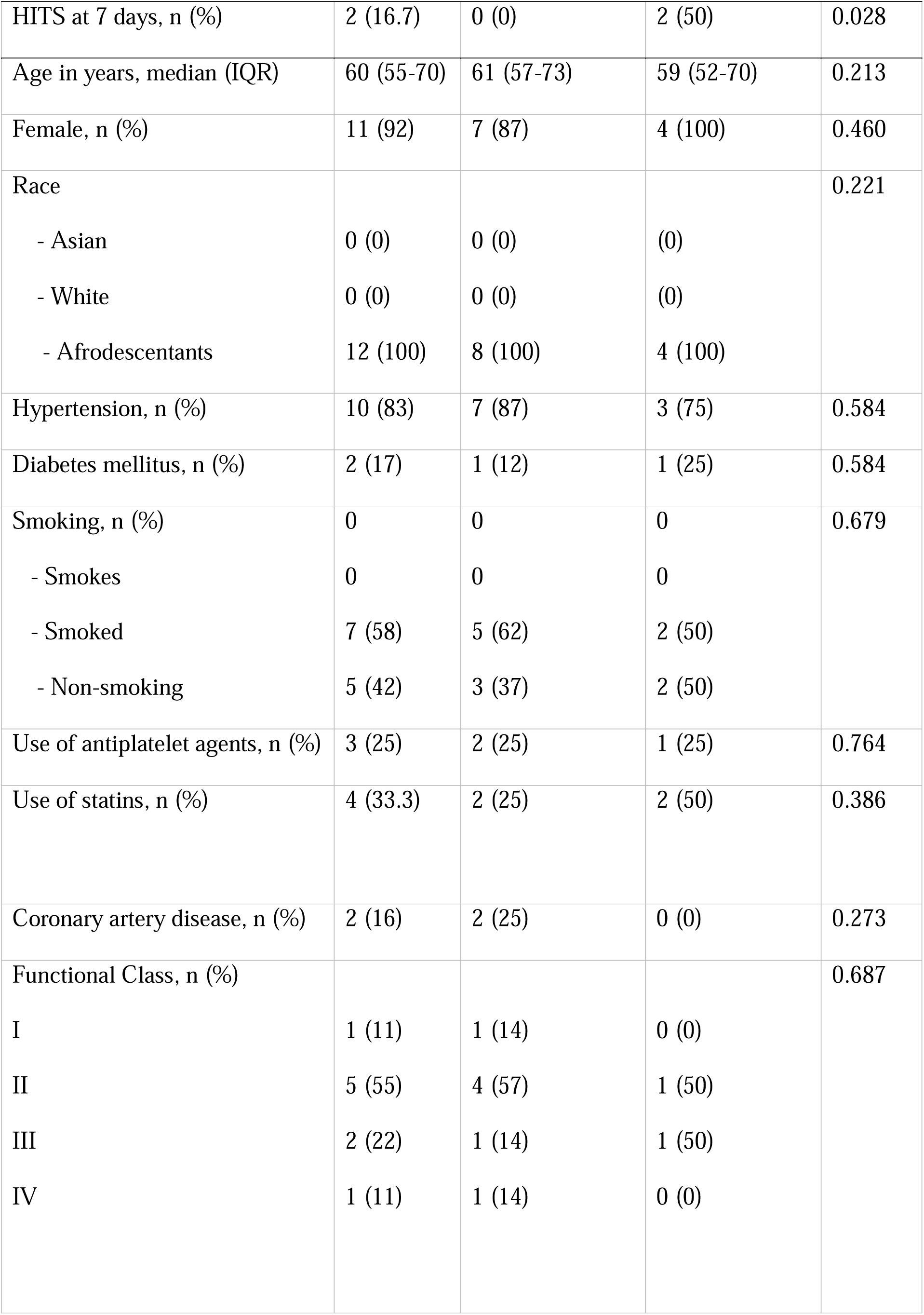
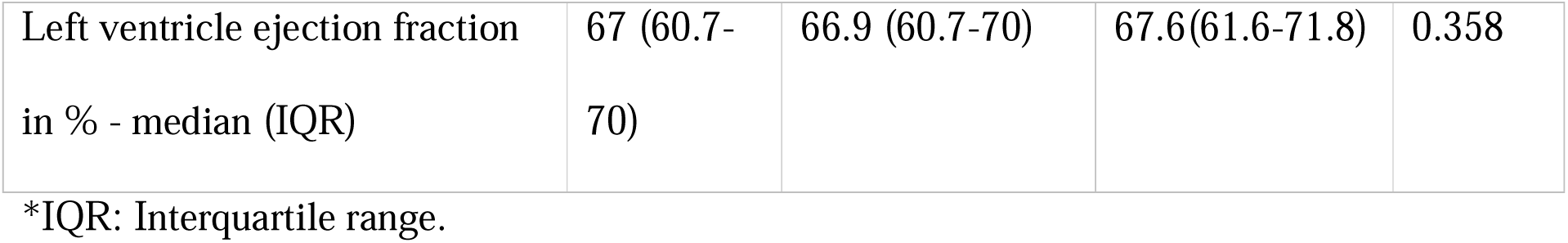
Primary outcome and clinical characteristics of Chagas disease patients randomized to acetylsalicylic acid (ASA) or control groups. HITS = high-intensity transient signals.

The median number of HITS decreased from 4.0 [(interquartile range (IQR) 1.3 – 6.8) to 0.5 (IQR 0.0 – 4.8) in the control group (within-group p-value=0.095); and from 3.5 (IQR 1.3 – 6.0) to none (in all patients) in the ASA-treated group (within-group p-value=0.012). However, the between-group difference was not significant (p=0.262). The Figure 1 shows the difference in number of HITS between days zero and seven. The figure 2 shows TCD control and intervention patients at day 0 and after 7 days.

**Figure 1:**
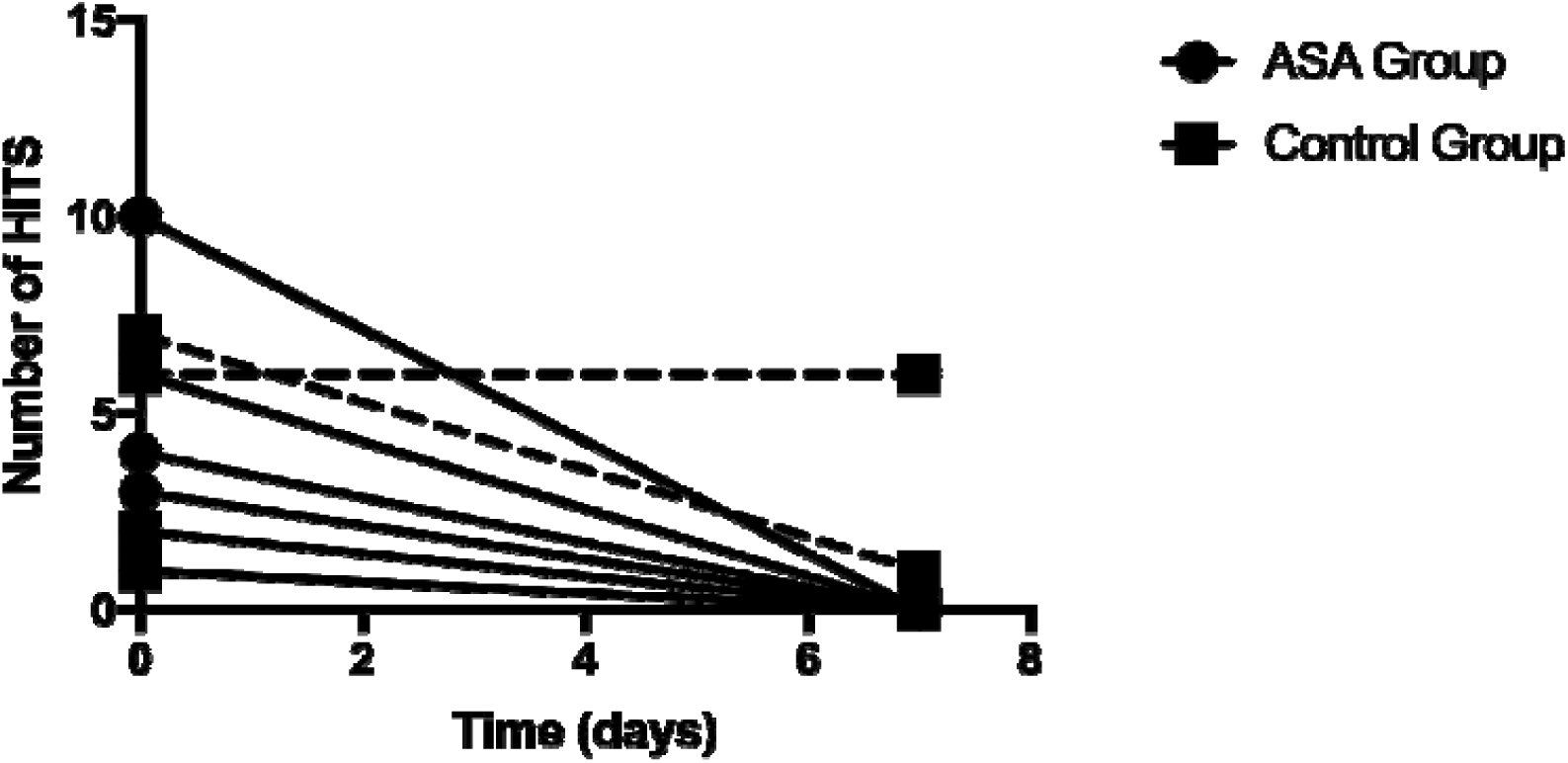
Number of high-intensity transient signals (HITS) at baseline (day 0) and 7 days in acetylsalicylic acid (ASA)-treated or control group patients.

**Figure 2.**
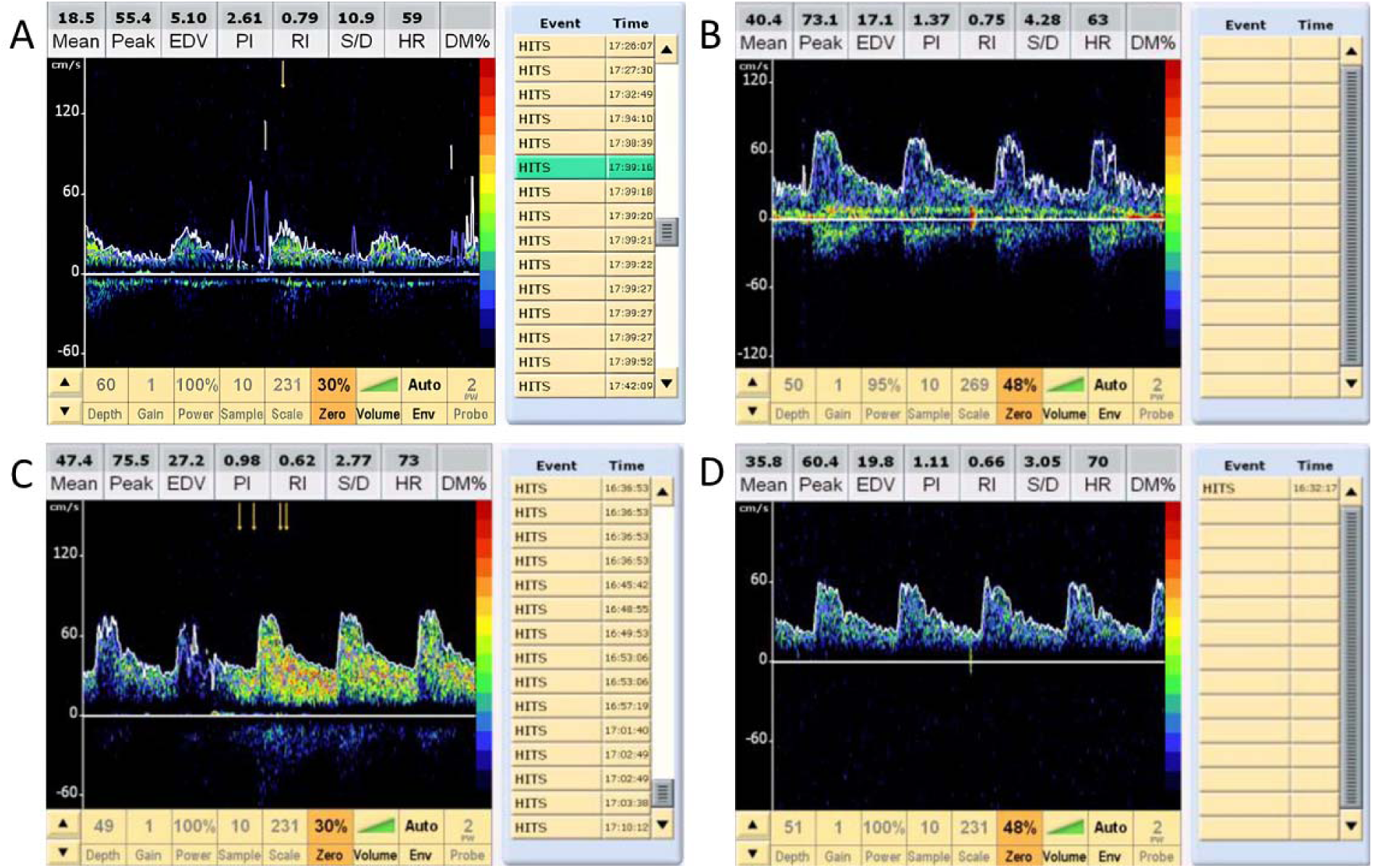
Patient in their 60s with Chagas disease randomized to acetylsalicylic acid use, LVEF= 66.9%. A: Baseline TCD showing HITS; B: TCD at 7 days showing no HITS. **Patient in their 60s with Chagas disease randomized to the control group, LVEF= 73%. C: Baseline TCD showing HITS; D: TCD at 7 days showing HITS.**

## Discussion

This proof-of-concept PROBE trial was carried out to evaluate the effectiveness of ASA in reducing the rate of cerebral microembolism in patients with chagasic HF. The main novel finding was a significant reduction in the proportion of HITS from baseline TCD after seven days of treatment with ASA 300mg in comparison with the control group. Additionally, Chagas patients with HITS treated with ASA showed a significant reduction in the number of HITS. Studies have demonstrated that antithrombotic therapy reduces the occurrence of microembolism in patients with carotid atherosclerotic disease, however the effectiveness of ASA in reducing the rate of HITS in patients with HF, being tested as primary prophylaxis, had not been demonstrated until now.^11, 12^ According to the Brazilian Guidelines on Antiplatelets and Anticoagulant Agents in Cardiology,^13^ warfarin is indicated for primary prevention of stroke in chagasic patients considered at high risk, such as those with left ventricular thrombus or aneurysm. However, one cohort study actually identified the use of ASA as risk factor for stroke in patients with heart failure, possibly due to a deleterious effect on cardiac function (Cerqueira-Silva et al., 2022).^2^ Since our study only evaluated short-term effects of ASA treatment, longer-term effects of ASA or other anti-thrombotics should be studied in Chagas disease before firm recommendations can be made.

Patients with chagasic cardiomyopathy presented with a numerically higher proportion of HITS when compared to non-chagasic patients, data that agrees with another study, but did not reach statistical significance.^7^ Moreover, in the multivariate analysis Chagas disease was not an independent risk factor for HITS in the first TCD. The other study^7^ investigated patients with higher baseline risk of HITS, such as those with a history of stroke, which we could not include because these patients are usually already using anti-thrombotic medications. It is possible that our study was underpowered to show a difference in microembolic burden in this lower-risk population.

Other reasons for a lower-than-expected embolic risk may exist for this patient sample. Patients were younger than most stroke cohorts and predominantly female^7, 14^ potentially lowering microembolic burden due to cardiovascular protective effects of endogenous estrogen in this age group. Patients with HF were predominantly mild/moderate functional class and frequently showed preserved left ventricular ejection fraction. Several studies have demonstrated a higher risk of embolism associated with reduced left ventricle ejection fraction, suggesting we selected a HF population with lower risk of embolism.^15, 16^ Chronic inflammation in Chagas disease results in endothelial dysfunction that can stimulate the hemostatic system, increasing fibrin production and platelet activation and impacting the formation of microemboli, but this was not enough to account for a statistically significant increase in HITS in our sample.^17–20^

The limitations of the study refer to the early interruption and sample size not being reached, due to the repercussions resulting from the pandemic and the long time of the study. Some patients from the original cohort also suffered a stroke or died during follow-up, making them unavailable for the present trial.

In conclusion, we did not replicate the finding of a higher rate of HITS in patients with Chagas disease when compared to non-Chagas patients. In the patients with Chagas disease where HITS were identified, seven days of ASA treatment decreased the proportion of HITS when compared to no ASA treatment. Long-term effects of primary ASA prophylaxis in high-risk populations for stroke in Chagas disease should be further studied.

## Non-standard Abbreviations and Acronyms

ASA: acetylsalicylic acid
ELISA: Enzyme-Linked Immunosorbent Assay
IQR: Interquartile range
LVEF: Left ventricle ejection fraction
HITS: High Intensity Transient Signals
HF: Heart Failure
MCA: middle cerebral arteries
PROBE: Prospective, randomized, open-label, blinded clinical
QVSFS: Questionnaire for Verifying Stroke-Free Status
TCD: Transcranial Doppler

## Data Availability

I declare the availability of all data referenced in the manuscript

## Acknowledgments

the Stroke Clinic, Hospital Professor Edgard Santos, Federal University of Bahia, Brazil.

